# A Wearable Thermoelectric Device for Closed-Loop Pulmonary Function Monitoring, Screening, and VR-Assisted Rehabilitation

**DOI:** 10.64898/2026.07.06.26357262

**Authors:** Lai Wei, Zheng Zhu, Xuan Zheng, Xinxin Yan, Han Tang, Chang Li, Zhaoyu Li, Yue Hou, Ziyu Wang

## Abstract

Early screening for chronic obstructive pulmonary disease (COPD) is critical due to the progressive and debilitating nature. Preliminary diagnosis typically relies on pulmonary function tests, particularly the ratio of forced expiratory volume in one second (FEV_1_) to forced vital capacity (FVC). However, conventional spirometers are often bulky and non-portable, while most existing portable devices can only measure a single parameter, such as FVC, thereby limiting comprehensive assessment. To address these limitations, an integrated wearable system was proposed for both monitoring and rehabilitation training. This system is based on the innovative thermoelectric-airflow inversion (TAI) model, which quantitatively correlates convective heat transfer with thermoelectric voltage to reconstruct airflow velocity and volume in real time. The developed thermoelectric smart mask enables simultaneous measurement of two key obstructive indicators (FVC and FEV_1_) and automatically evaluates COPD risk via the FEV_1_/FVC ratio, alerting users to seek medical consultation when abnormalities are detected. In terms of performance, the device demonstrates a measurement accuracy of 99.10% and a coefficient of determination (R^2^) of 0.9947 compared to a commercial spirometer. Furthermore, the incorporated virtual reality assisted rehabilitation system was developed, yielding an average FVC improvement of 5.87% across three participants after one week of interactive training. Enabled by the TAI framework and a closed-loop multi-parameter design, this platform provides an intelligent, quantitative, and continuous solution for respiratory healthcare and rehabilitation.

## 1. Introduction

Chronic obstructive pulmonary disease (COPD) is a major cause of morbidity and mortality worldwide, posing an escalating public health challenge driven by aging populations and persistent exposure to risk factors such as smoking and air pollution[1–3]. Early detection and timely intervention are critical for improving patient outcomes and alleviating the associated healthcare burden.[4–8] Pulmonary function testing remains the gold standard for diagnosing COPD, with diagnosis primarily based on spirometric parameters including forced vital capacity (FVC) and, more importantly, forced expiratory volume in one second (FEV_1_)[9–13]. However, conventional spirometers are large, costly, and require operation by trained personnel, rendering them impractical for community based or home based screening.[14] These limitations underscore the urgent need for portable, low-cost, and user-friendly devices capable of accurately measuring FEV_1_ and FVC in non-clinical settings.[15]

Recent years have witnessed significant progress in the development of portable and wearable respiratory monitoring technologies, driven by advances in flexible electronics, microelectromechanical systems, and wireless sensing[16–18]. Various transduction mechanisms, such as pressure-based differential flow sensors[19], hot-wire or thermistor-based thermal anemometers[20,21], and piezoresistive or capacitive strain sensors for chest-wall motion[22], have been explored for estimating airflow or breathing patterns. Some low-cost flowmeters and mobile devices can estimate FVC[23–30], yet few can accurately capture both FVC and FEV_1_ with medical-grade precision. Such dual-parameter accuracy is essential for early COPD screening and disease monitoring[12]. Consequently, the development of a wearable and dual-parameter respiratory sensor, typically the FEV_1_ and FVC, remains an unmet challenge.

Thermal-based flow sensing has long been recognized for its simplicity, fast response, and reliability in measuring gas velocity[31–36]. Owing to their high temperature sensitivity and solid-state configuration, thermoelectric devices (TED) have been widely employed in respiration or airflow detection[37–39]. The Seebeck effect enables them to directly convert temperature gradients into voltage signals, offering self-powered potential and robust environmental adaptability[40]. A previous study systematically investigated the physical mechanisms underlying the airflow-thermoelectric response relationship, and quantitatively revealed a deterministic relationship between airflow velocity and the output voltage of thermoelectric elements under controlled conditions[33]. This earlier study, along with other research efforts, has successfully applied thermoelectric technology to microenergy harvesting[41,42], high-precision temperature sensing[43], and environmental or low-velocity flow monitoring[44,45]. However, the absence of a quantitative theoretical framework capable of reconstructing airflow velocity from voltage signals severely limits its application in quantitative medical diagnostics and closed-loop respiratory training and rehabilitation.

Most existing studies, including prior work focusing on the signal generation mechanism, have primarily concentrated on the forward problem of how airflow perturbations generate electrical signals. However, for spirometric diagnosis and rehabilitation guidance, quantification of specific respiratory parameters is required, which necessitates solving an inverse problem: reconstructing airflow velocity (and subsequently volumetric flow rate) from the voltage response under well-defined boundary conditions. Consequently, most reported thermoelectric sensors serve only as empirical indicators of relative airflow intensity rather than quantitative spirometric instruments. This limitation stems primarily from the undefined boundary conditions in open environments, which render the inverse problem—reconstructing absolute velocity from voltage signals—mathematically intractable. To address this, we developed a confined flow environment that physically regularizes the near-surface laminar flow profile and defines an effective cross-sectional area, thereby establishing the necessary boundary conditions for physics-based inversion and enabling the reconstruction of absolute pulmonary parameters.

To address this gap, the Thermoelectric-Airflow Inversion (TAI) model was proposed, which quantitatively establishes the relationship between convective heat transfer and thermoelectric voltage under laminar flow conditions. By integrating Newton’s law of cooling with the Nusselt-Reynolds correlations for exploring the relationship between airflow velocity and Seebeck-induced voltage, the analytical model is established for real-time reconstruction of pulmonary flow and volume. Based on the TAI framework, a wearable TED-based respiratory sensing system is developed, capable of real-time, dual-parameter measurement of FVC and FEV_1_ for early COPD screening. We developed a lightweight smart mask integrated with a custom-built TED to capture airflow dynamics. The generated electrical signals are transmitted to a host computer for computational processing, enabling real-time flow reconstruction for respiratory diagnosis. Experimental evaluations were conducted using healthy adult volunteers, simulated obstructive cases, and de-identified clinical participants with non-COPD cardiopulmonary conditions. Compared with a commercial spirometer, our results demonstrate accurate spirometry of FEV_1_ and FVC, with an accuracy of 99.10% and a coefficient of determination (R²) of 0.9947. This capability further enables COPD-oriented risk stratification, as the reconstructed FEV_1_/FVC ratio and FVC values correctly separated all simulated cases.

For individuals whose FVC falls below the lower limit of normal (LLN), the system activates a targeted virtual reality (VR)-assisted rehabilitation module, forming a functional closed-loop pathway from quantitative assessment to condition-triggered intervention. Following one week of VR-assisted training, participants with baseline FVC values below the LLN exhibited progressive improvements in pulmonary function, with an average FVC increase of 5.87%, demonstrating the feasibility of condition-triggered closed-loop rehabilitation.

## 2. Results and Discussion

### 2.1. Closed-Loop Respiratory Framework and System Overview

The proposed system establishes a closed-loop respiratory healthcare framework that integrates wearable thermoelectric sensing, quantitative pulmonary function assessment, automated COPD risk screening, and VR-assisted rehabilitation within a single platform (Figure 1). As illustrated in Figure 1a, the system is implemented through a lightweight, 3D-printed smart mask that integrates the TED and its associated control electronics. The mask incorporates a confined airflow channel designed to geometrically regulate the expiratory flow field, ensuring well-defined boundary conditions for accurate airflow sensing. To compare the performance and functional positioning of this wearable platform, Figure 1b presents a radar chart comparing the proposed system with representative state-of-the-art respiratory monitoring technologies.[23,25,26,29,30] The comparison evaluates 6 key dimensions relevant to practical pulmonary assessment: closed-loop capability, wearability, functionality of direct measurement, measurable parameters, environmental interference immunity, and correlation (R²) with measurements from a commercial spirometer. Consistent scoring criteria were applied across all studies. This comparison highlights that the proposed system uniquely integrates multi-parameter spirometry, high quantitative accuracy, and closed-loop functionality within a single wearable device.

**Figure 1.**
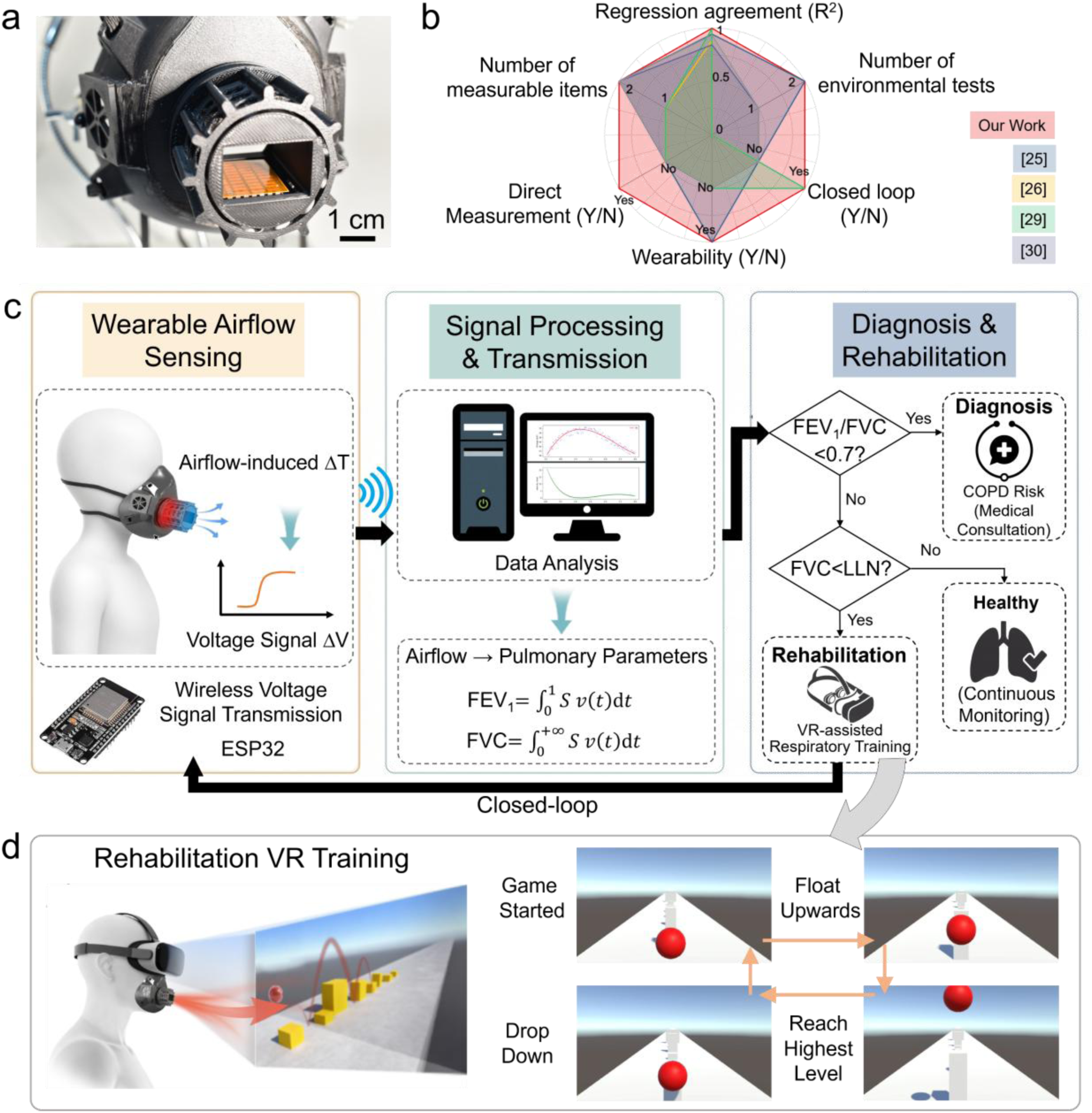
Overview of the closed-loop thermoelectric respiratory assessment framework. (a) Photograph of the lightweight 3D-printed thermoelectric smart mask. (b) Radar chart comparing R^2^, closed-loop capability, wearability, number of environmental tests, direct measurement, and number of measurable parameters with advanced works. (c) Schematic diagram of the workflow integrating thermoelectric sensing, COPD risk analysis, and VR-based rehabilitation feedback. (d) VR training paradigm driven by reconstructed expiratory airflow velocity.

Figure 1c illustrates the closed-loop operational workflow of the system. During exhalation, airflow across the upper surface of the TED induces convective heat transfer, generating a voltage signal (ΔV). This signal is wirelessly transmitted to the host computer via an ESP32 module, where the TAI-based algorithm reconstructs real-time airflow velocity and computes pulmonary parameters, including FEV_1_ and FVC. Based on these quantitative outputs, the system automatically assesses COPD risk using the FEV_1_/FVC ratio: a value below 0.7 triggers a screening alert and recommends medical consultation. If FVC falls below the lower limit of normal (LLN), the system activates a VR-assisted respiratory training module, thereby forming a continuous closed loop from quantitative assessment to condition-triggered rehabilitation.

Figure 1d details the VR-assisted rehabilitation mechanism. The reconstructed expiratory airflow velocity is used to control the motion of a virtual ball within an immersive virtual environment, providing intuitive biofeedback and enabling targeted respiratory training. This interactive paradigm not only engages the user but also reinforces proper breathing patterns through real-time visual guidance.

### 2.2. Airflow and stability testing

The airflow response and environmental stability of the TED were evaluated through Seebeck characterization, airflow-response measurements, and temperature-dependent testing (Figure 2). As illustrated in Figure 2a, forced airflow enhances convective heat transfer at the TED surface, thereby establishing the temperature difference across the thermoelectric legs and generating a measurable Seebeck voltage output. The intrinsic Seebeck behavior was first examined by applying temperature differences ranging from 0 to 20 K across the TED. Here, Δ*T* = *T*_upper_ − *T*_lower_ denotes the temperature difference between the upper (air-exposed) and the lower surface of the TED. Representative voltages at ΔT = 2.9, 9.8 and 20 K, are 8.5, 31.7 and 65.5 mV respectively, as shown in Figure 2b, confirming the robustness and repeatability of the developed module.

**Figure 2.**
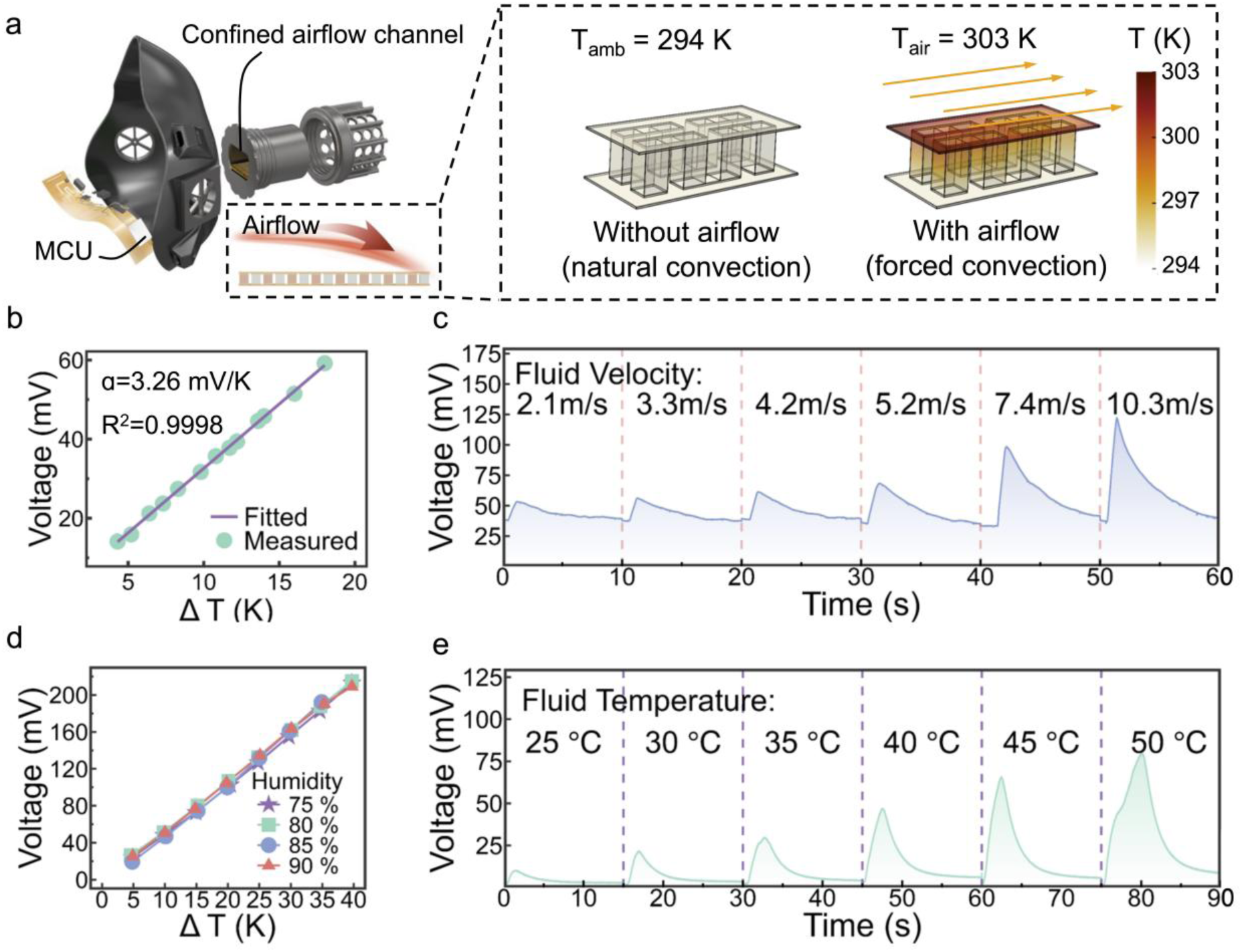
Thermoelectric sensing mechanism and environmental performance characterization. (a) Schematic illustrating the airflow-induced convective heat transfer on the TED. (b) Seebeck response under different temperature differences. (c) Voltage response under different airflow velocities. (d) Voltage-temperature-difference curves under varying humidity. (e) Voltage response at different fluid temperatures.

To assess flow sensitivity, the device was exposed to controlled laminar airflows of varying velocity (2.1-10.3 m/s), and the output voltage increased with flow speed (Figure 2c), consistent with the TAI-predicted convective cooling mechanism. Previous physiological studies and fluid-dynamic analyses suggest that, within the confined geometry of a mask, the effective airflow velocity impinging on the sensor typically ranges from approximately 0.9 to 6 m/s, within the range of velocities tested in this work[46]. This trend confirms that airflow enhances convective heat transfer at the hot side, thereby enlarging the temperature gradient across the TED. To evaluate thermal stability, experiments were repeated with airflow temperatures ranging from 25 to 50 °C. Higher temperatures yielded steeper voltage-time slopes and higher peak voltages (Figure 2e), consistent with the increased driving temperature difference between the airflow and TED surface.

The humidity-dependent stability of the TED was systematically evaluated by characterizing the voltage response as a function of temperature difference under controlled humidity conditions. As shown in Figure 2d, the V-ΔT curves exhibit a linear relationship across a range of humidity levels indicating that device transduction remains robust against ambient moisture variations. These results collectively demonstrate the stable performance of the proposed device under diverse environmental and operational conditions.

### 2.3. TAI Model and Numerical Verification

Considering convective heat transfer with Seebeck-induced voltage generation, the TAI model was established to quantitatively describe the thermoelectric response under laminar airflow, and the detailed mathematical derivation is provided in the Supporting Information (Section S2, Equations S1-S10). When warm airflow passes over the surface of the TED, a thin thermal boundary layer forms at the solid-fluid interface. Under confined airflow conditions in the mask’s channel, according to the classical boundary-layer theory, a laminar boundary layer is formed as a no-slip condition at the solid surface, resulting in a stable and quasi-steady velocity profile adjacent to the device surface.[47] Through forced convection, the heat is transferred from the gas to the upper layer of the thermoelectric leg, and the absorbed energy is then conducted through the thermoelectric couples within the TED, and thus converted into an electric potential difference via the Seebeck effect. The rate of heat exchange at the interface follows Newton’s law of cooling, which can be expressed as:

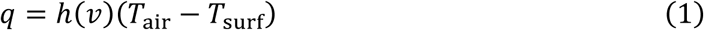

where the convective heat-transfer coefficient ℎ(*v*) increases with flow velocity following the Nusselt-Reynolds correlation. The heat transfer in the fluid domain and solid domain, together generates a temperature difference across the TED, which produces an exponentially rising voltage signal, as schematically illustrated in Figure 3a.

**Figure 3.**
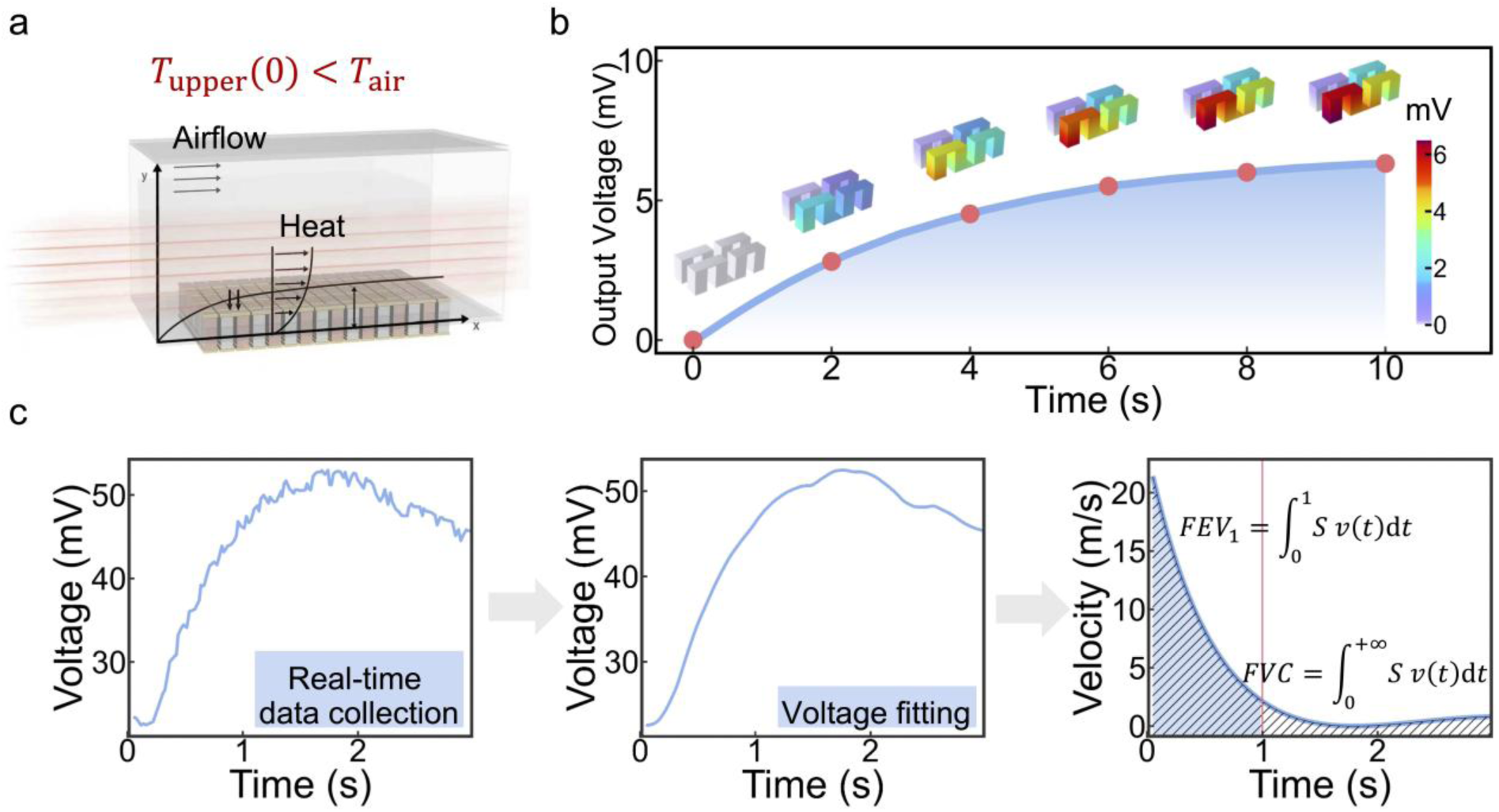
TAI model and numerical verification. (a) Schematic of convective heat transfer between laminar airflow and the thermoelectric device surface. (b) COMSOL simulated the exponential voltage-time response and the internal electric potential distribution. (c) Data-processing pipeline: voltage acquisition, curve fitting, velocity inversion and flow-volume integration.

Based on the previous study that established a forward model for predicting the voltage difference as a function of flow velocity^[34]^, we here derive the corresponding inverse solution to reconstruct flow velocity from the measured voltage signal. The exhaled volume is then obtained through numerical integration of the velocity over the exhalation period.

According to the Seebeck effect, the thermoelectric voltage follows

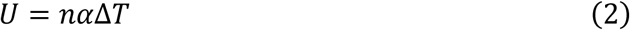

where *n* is the number of thermoelectric couples, *α* is the Seebeck coefficient, and Δ*T* is the temperature difference across the TED. Combining Newton’s law of cooling with the Nusselt-Reynolds correlation, the forced laminar convection yields an exponential voltage-time response^[34]^:

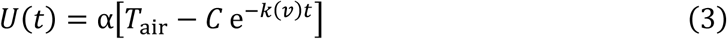

where *C* = *T*_air_ − *T*_upper_(0), and *k*(*v*) is a velocity-dependent relaxation coefficient. Solving for *U*(*t*) yields the following expression for *v*(*t*) gives

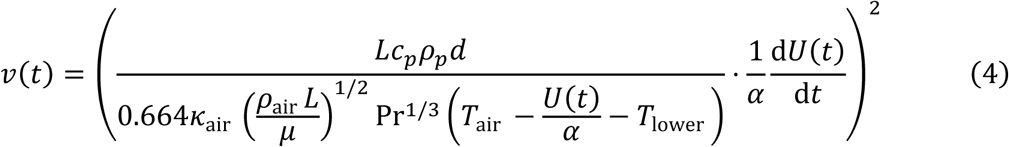

where *L*, *c*_*p*_, *ρ*_*p*_, and *d* are the characteristic length, specific heat, density, and thickness of the TED, respectively. The volumetric flow is then obtained by numerical integration:

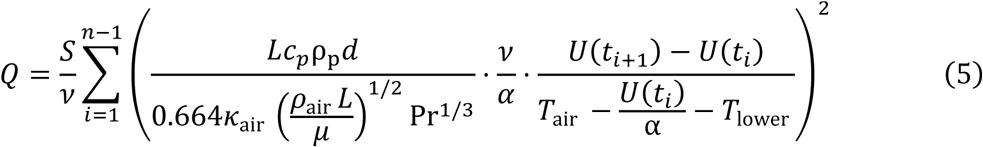

where *S* is the effective cross-sectional area and *v* is the sampling rate.

To validate the proposed analytical model, COMSOL Multiphysics simulations were performed under the same conditions as the physical experiment: an airflow velocity of 3 m/s, an initial ΔT of 8 K between airflow and device, and a TED composed of four arranged P-N pairs. The simulated open-circuit voltage *U*(*t*) exhibits an exponential trend: within 10 seconds, the voltage rises from 0 to 6.32 mV as heat transfer progresses and then approaches a steady state (Figure 3b), consistent with the proposed TAI-predicted temporal dynamics of the thermoelectric response. The TAI framework was further verified and implemented through a numerical-experimental workflow that bridges theory and practice (Figure 3c). The data-processing pipeline includes real-time voltage acquisition, curve fitting, and velocity calculation through the TAI model, followed by numerical integration to compute pulmonary parameters such as FEV_1_ and FVC. These results confirm that the TAI framework accurately captures the transient thermoelectric response, and provide a computational model for reconstructing expiratory airflow velocity and the exhaled volume.

### 2.4. Airflow Measurement and Agreement Analysis

The quantitative airflow-sensing performance of the TED was evaluated by comparing experimentally recorded voltage-time responses with the result predicted by the TAI model (Figure 4a-c). At representative velocities of 2.2, 3.3, and 3.9 m/s, the measured voltage curves closely matched both the theoretical, with all R^2^ exceeding 0.9984. These results confirm that the thermoelectric voltage response under forced convection follow an exponential form, as predicted by the TAI model.

**Figure 4.**
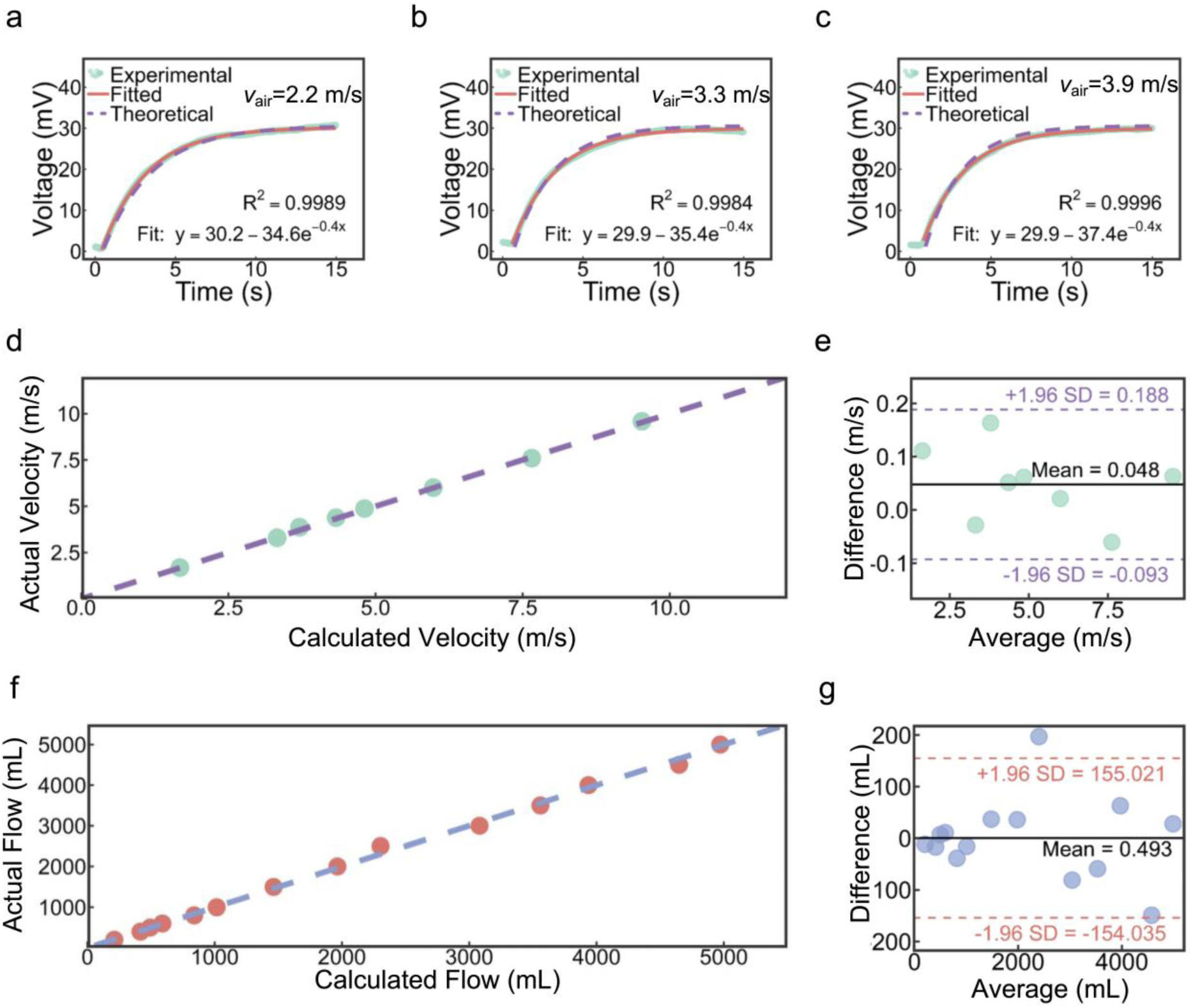
Airflow measurement and agreement analysis. Experimental voltage-time signals and fitted exponential curves derived from the TAI equations at different flow velocities: (a) 2.2 m/s, (b) 3.3 m/s, and (c) 3.9 m/s. Comparison between calculated and measured airflow velocities (d), and the corresponding BA analysis (e). Comparison between calculated and measured flow (f), and the corresponding BA analysis (g).

Based on the fitted voltage responses, instantaneous airflow velocities reconstructed via the TAI model were compared with reference values obtained from a digital anemometer. Across eight airflow conditions spanning 2-10 m/s, the reconstructed velocities showed strong agreement with the measured values, with no evident over- or underestimation trend (Figure 4d). Then, Bland-Altman (BA) analysis[48,49] performed to quantify agreement between predicted and measured results (Figure 4e), as the mean bias is 0.0479 m/s, and the 95% limits of agreement (LoA) range from −0.0926 to +0.1884 m/s. All eight data points fell within these limits, and the differences exhibited no dependence on the magnitude of velocity, suggesting the absence of proportional bias. Therefore, these results demonstrate that the TAI model enables accurate reconstruction of airflow velocity across the full range of tested conditions.

Next, the volumetric flow can be obtained by integrating the reconstructed velocity over time. Comparison with real measured values, the predicted ones demonstrate excellent agreement throughout the 14 expiration-like flow conditions (Figure 4f). The BA analysis further confirms the effectiveness of the proposed method, as the mean bias is 0.493 mL, and the LoA ranges from −154.04 to +155.02 mL (Figure 4g). Most of the data points (92.9%) fell within the LoA, and no proportional bias is observed across the full test range of 0-5000 mL. The above analysis shows that the proposed TAI-based reconstruction can provide clinically relevant accuracy for volumetric flow estimation under controlled flow conditions.

### 2.5. VR-Assisted Pulmonary Training and Closed-Loop Feedback

The wearable thermoelectric sensing system was integrated into a lightweight 3D-printed smart mask for spirometric assessment and COPD-oriented screening. The mask incorporates a flexible printed circuit board (FPCB) that serves as the electrode layer, onto which the TED is mounted on its inner surface. This configuration exposes the TED directly to the exhaled airflow, enabling sensitive thermal exchange. The complete system, comprising the TED array, signal acquisition circuit, and wireless transmission module, weighs less than 100 g and introduces negligible breathing resistance, supporting its potential use as a wearable spirometric assessment platform.

To verify quantitative accuracy under human use, FVC was measured using both the TAI-based wearable system and a commercial spirometer across fourteen de-identified measurements (Figure 5a). The validation cohort included de-identified measurements from healthy adult volunteers, simulated obstructive cases generated using a breathing simulator, and clinical participants with non-COPD cardiopulmonary conditions. Each participant performed five spirometric maneuvers following standard forced-exhalation procedures, and the averaged values were used for inter-device comparison. The FVC measurements obtained by the proposed system showed excellent agreement with those from the commercial handheld spirometer (NaiLi model), achieving a coefficient of determination R² of 0.9947 and a mean relative accuracy of 99.10%. The BA analysis further confirmed excellent agreement between the two methods (Figure 5b): the mean bias was 8.4 mL, with a standard deviation of 50.38 mL and the 95% LoA ranging from −90.31 to +107.17 mL. All fourteen data points fell within these limits, and no proportional bias was observed, indicating a high level of agreement between the wearable thermoelectric system and the reference spirometer for FVC quantification.

**Figure 5.**
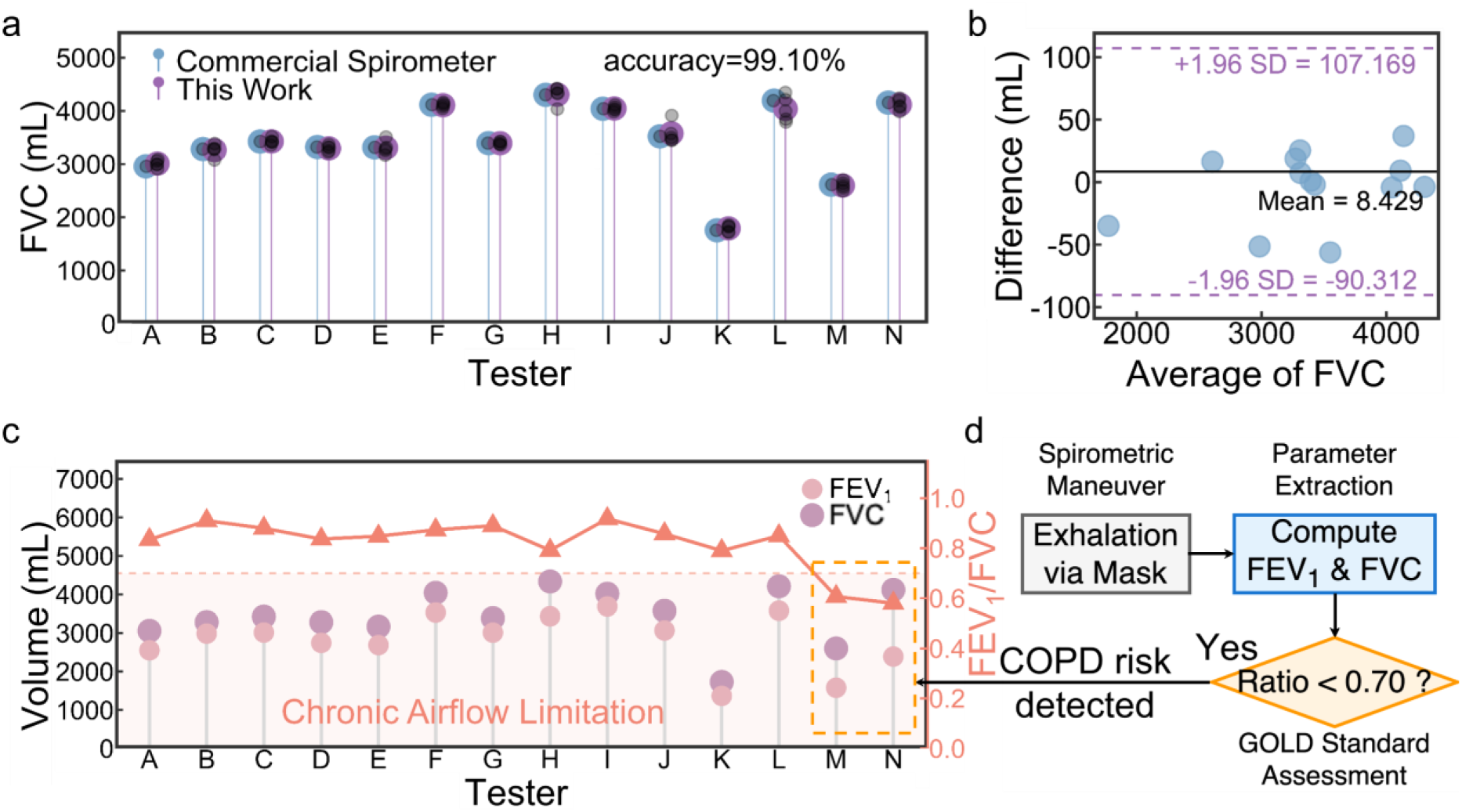
Wearable thermoelectric spirometry and COPD-oriented diagnostic validation. (a) Comparison of FVC measured by this work and a commercial spirometer. (b) BA analysis of FVC measurement agreement. (c) Measured FEV_1_ and FVC values for 14 measurements. (d) Diagnostic decision flow illustrating COPD risk identification based on the FEV_1_/FVC ratio.

Beyond FVC, the system can simultaneously extract FEV_1_ for each measurement, enabling identification of obstructive airflow patterns based on the Global Initiative for Chronic Obstructive Lung Disease (GOLD) spirometric criterion (Figure 5c, d). In this cohort, measurements from healthy volunteers and non-COPD clinical participants exhibited FEV_1_/FVC ratios ranging from 0.79 to 0.92. In contrast, the two simulated obstructive cases showed markedly reduced ratios of 0.58–0.63, falling below the GOLD threshold of 0.70 (Figure 5c). These results demonstrate the system’s capability to identify obstructive airflow patterns via the FEV_1_/FVC ratio.

To enable assistive therapy and respiratory rehabilitation, a VR-assisted pulmonary training module that leverages real time reconstructed airflow velocity to provide interactive, breathing-dependent visual feedback was developed (Figure 6a, b). In this module, the reconstructed airflow velocity is mapped to the vertical motion of a virtual ball, allowing users to perform goal-oriented training task in a VR environment. The designs aim to actively motivate users to achieve and sustain a predefined breathing target, rather than passively observe visual feedback.

**Figure 6.**
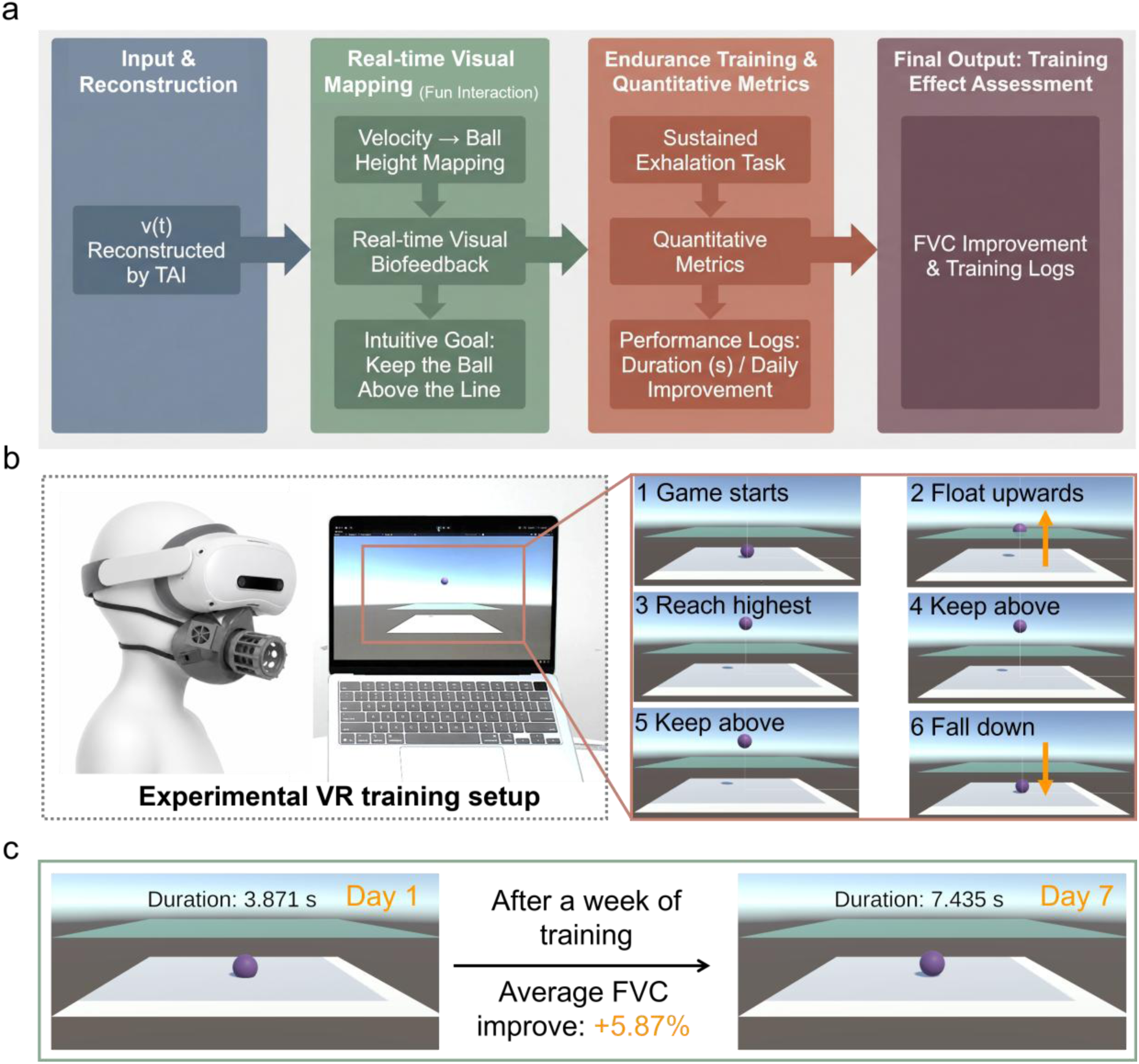
VR-assisted closed-loop respiratory training enabled by thermoelectric airflow reconstruction. (a) Schematic of the closed-loop VR training principle. (b) The experimental VR training setup and representative screenshots of the VR task. (c) The result of repeated VR training tasks.

Specifically, higher expiratory airflow velocities propel the ball to greater heights, directly linking respiratory effort to task progression. When the instantaneous velocity exceeds a predefined threshold, the ball rises above a reference plane; otherwise, it descends below it. Users are instructed to maintain the ball above this plane for as long as possible, thereby transforming sustained exhalation into a clear and intuitive training objective.

The duration for which the virtual ball remains continuously above the reference plane serves as a quantitative metric of expiratory endurance, breath stability, and voluntary airflow control. By embedding quantitative airflow feedback within an interactive, game-like task, the VR module enhances user engagement and motivation during repetitive respiratory training. Representative training outcomes are shown in Figure 6c, with detailed daily results provided in Figure S7.

To evaluate the feasibility of VR-assisted closed-loop rehabilitation, multiple participants performed daily VR-based respiratory training sessions for seven consecutive days. Across all three participants, consistent improvements were observed in both behavioral performance and spirometric outcomes. FVC increased from baseline by an average of 5.87% after seven consecutive days of training (Figure S7). This improvement was accompanied by a progressive increase in the VR task duration metric, reflecting enhanced expiratory endurance and breathing stability.

Participants with baseline FVC values were below or close to the LLN, as defined by the GLI-2012 reference equations[50], exhibited gradual increases in FVC over the training period. After repeated sessions, their values approached or exceeded the corresponding LLN thresholds (Figure S7). These results demonstrate that an LLN-triggered, interactive VR module can integrate game-based engagement with quantitative respiratory targets, enabling coordinated improvements in both functional breathing performance and spirometric capacity.

## 3. Conclusion

This work presents a wearable thermoelectric respiratory sensing system that integrates quantitative pulmonary assessment with VR-assisted rehabilitation within a unified closed-loop framework. The proposed TAI model establishes a physics-based relationship between convective heat transfer and thermoelectric voltage under confined laminar airflow conditions, enabling real-time reconstruction of airflow velocity and exhaled volume. Systematic validation across 14 participants, including simulated COPD subjects and real patients with cardiopulmonary diseases, demonstrates excellent agreement with commercial spirometry (R^2^=0.9947, 99.10% accuracy relative to reference) and the capability to distinguish normal from obstructive respiratory patterns. Integrated with the VR-assisted feedback module, the system achieved an average FVC improvement by 5.87% across three participants after one week of training, supporting its potential for adaptive pulmonary rehabilitation. By combining a physically grounded theoretical model, human-validated performance, and interactive feedback, this study introduces a novel thermoelectric-sensing-driven paradigm for intelligent, low-cost, and continuous respiratory healthcare. This framework lays the foundation for future quantitative, home-based COPD monitoring and rehabilitation across diverse respiratory conditions.

## 4. Materials and Methods

### 4.1. Device Fabrication

The TED was assembled using commercially thermoelectric legs with dimensions of 1.4×1.4×2.5 mm^3^. The patterned copper electrodes are printed on polyimide (PI) film, which is used as FPCB to form the hot or cold sides of the module. 70 P-N thermoelectric legs were alternately soldered in a 10×7 matrix, and sandwiched by two FPCBs. Electrical connections between adjacent P-N elements were established directly through the copper traces on the FPCB without additional bonding materials or encapsulation layers. The assembled TED had an open structure without surface encapsulation to ensure efficient convective heat. Crucially, the encompassing airflow channel allows the sensor to operate under well-developed and stable convective flow conditions. The developed channel structure in the mask eliminates external environmental disturbances and enforces a no-slip condition at the walls[47], creating a stable, quasi-steady laminar velocity profile essential for precise velocity inversion.

### 4.2. Measurement Setup

To evaluate the thermoelectric response under controlled airflow conditions, the TED was mounted on a custom test fixture that directed laminar airflow across its upper surface. Airflow was generated using a fan, and the velocity was independently verified by a digital anemometer (UT363, Uni-T, China). This configuration allowed continuous adjustment of flow speed while maintaining a stable airflow direction.

The output voltage of the TED was acquired using a 16-bit ADS1115 analog-to-digital converter interfaced with an ESP32 microcontroller. The sampling rate was set to 100 Hz to capture the transient voltage response. The ESP32 transmitted the acquired data via Wi-Fi to a host computer for real time visualization and processing. A Python program was developed to receive the data stream, perform curve fitting, and extract FEV_1_ and FVC parameters according to the TAI model. This setup enabled real time quantitative airflow reconstruction and spirometric parameter extraction. The wiring diagram of the complete acquisition system (ADS1115-ESP32-TED) is provided in Figure S6 of the Supporting Information.

### 4.3. Human Testing Protocol

A total of fourteen measurements were analyzed, including ten healthy volunteers, two simulated COPD subjects, and two real patients with cardiopulmonary diseases. These cases respectively cover the normal, obstructive respiratory patterns, and other clinically relevant symptoms. The healthy participants had no known respiratory disorders, while the simulated COPD subjects were generated using a breathing simulator to replicate reduced FEV_1_/FVC ratios (<0.7), mimicking the airflow obstruction characteristic.

Human respiratory measurements were conducted in accordance with ethical standards approved by the institutional ethics committee. Each participant was instructed to don the smart mask and, following a full inspiration to total lung capacity, perform five consecutive forced exhalation maneuvers in accordance with the ATS/ERS standard spirometry protocol. The study protocol adhered to Declaration of Helsinki and received approval from the ethics committee of Zhongnan Hospital of Wuhan University (2023046K). Informed consent was obtained from patients and volunteers before study. The voltage signals were continuously recorded during each exhalation and processed in real time through the TAI-based algorithm to compute FEV_1_, FVC, and the FEV_1_/FVC ratio. For validation, measurements were simultaneously taken using a commercial handheld spirometer (NaiLi electronic spirometer, China) under identical test conditions. Agreement between the two measurement methods was assessed using linear regression analysis (R^2^) and BA analysis.

A pilot VR-assisted pulmonary training experiment was further conducted to assess the feasibility of the proposed closed-loop rehabilitation strategy, in which multiple participants performed daily VR-based breathing sessions for seven consecutive days. The system recorded each session and re-evaluated the participant’s FVC after each day of training. Across the three participants, FVC exhibited a progressive increase after one week of VR-assisted training sessions, confirming the potential of integrating thermoelectric sensing with interactive VR feedback for pulmonary function improvement.

### 4.4. VR Module Design and Software Implementation

The VR training module was developed using the Unity 3D engine to create an interactive environment that responds to the user’s respiratory airflow in real time. The thermoelectric sensor signal, acquired by the ESP32 and transmitted to the host computer, was converted into instantaneous airflow velocity values via the TAI algorithm implemented in Python. These values were transmitted to the Unity interface, where they were mapped to the motion of a virtual object (a rising ball) to provide visual feedback during breathing exercises.

The VR environment was displayed using a Pico headset, enabling immersive visualization and intuitive user interaction. The training task required participants to maintain steady exhalation in order to keep the virtual ball above a target height, with the goal of enhancing respiratory endurance and voluntary airflow control. All gameplay data were recorded for analysis, including exhalation duration, airflow velocity, virtual object displacement, and the measured pulmonary parameters (FEV_1_, FVC). This closed-loop integration of thermoelectric sensing, algorithmic computation, and interactive VR feedback establishes a novel unified platform for both pulmonary assessment and rehabilitation.

## Data Availability Statement

The data supporting this article have been included as part of the Supplementary Information.

## Declaration of Generative AI and AI-Assisted Figure Preparation

Generative AI-assisted tools were used solely to create or refine schematic illustrations in Figures 1c and 6b. These schematic elements are illustrative and do not depict actual study participants. No generative AI tool was used to generate, modify, fabricate, or analyze experimental data, clinical data, or quantitative plots. All AI-assisted visual elements were reviewed and edited by the authors, who take full responsibility for the accuracy, integrity, and originality of the manuscript.

## Funding Sources

National Natural Science Foundation of China (Grant No. 12302220), the Translational Medicine and Interdisciplinary Research Joint Fund of Zhongnan Hospital of Wuhan University (Grant No. ZNJC202424) and Institute for Goneo New Energy Foundation of Wuhan University.

The authors sincerely thank Prof. Zhengyou Liu for providing the authorization to use COMSOL software, which has been essential for our research simulations.

